# Meat consumption and risk of 25 common conditions: outcome-wide analyses in 475,000 men and women in the UK Biobank study

**DOI:** 10.1101/2020.05.04.20085225

**Authors:** Keren Papier, Georgina K Fensom, Anika Knuppel, Paul N Appleby, Tammy YN Tong, Julie A Schmidt, Ruth C Travis, Timothy J Key, Aurora Perez-Cornago

## Abstract

**Background:** There is limited prospective evidence on the association between meat consumption and many common, non-cancerous health outcomes. We examined associations of meat intake with risk of 25 common conditions (other than cancer).

**Methods:** We used data from 474 998 middle-aged men and women recruited into the UK Biobank study between 2006 and 2010 and followed-up until 2017 (mean follow-up of 8.0 years) with available information on meat intake at baseline (collected via touchscreen questionnaire), and linked hospital admissions and mortality data. For a large sub-sample, dietary intakes were re-measured using an online, 24-hour questionnaire.

**Results:** In multi-variable adjusted (including body mass index (BMI)) Cox regression models corrected for multiple testing, a higher consumption of red and processed meat combined was associated with higher risks of ischaemic heart disease (HR per 70 g/day higher intake 1.14, 95% CI 1.06-1.22), pneumonia (1.28,1.15-1.41), diverticular disease (1.18,1.10-1.26), colon polyps (1.09,1.04-1.13), and diabetes (1.29,1.19-1.40), and a lower risk of iron deficiency anaemia (IDA), driven by a higher consumption of red meat (HR per 50g/day higher intake 0.77,0.69-0.86). Higher poultry meat intake was associated with higher risks of gastro-oesophageal reflux disease (HR per 30g/day higher intake 1.14, 1.06-1.23), gastritis and duodenitis (1.10,1.04-1.16), diverticular disease (1.09,1.04-1.16), and diabetes (1.13,1.06-1.20), and a lower risk of IDA (0.80,0.73-0.87).

**Conclusions:** Higher red, processed, and poultry meat consumption was associated with higher risks of several common conditions; higher BMI accounted for a substantial proportion of these increased risks. Higher red and poultry meat consumption was associated with lower IDA risk.

## Introduction

The World Health Organization^1^ and many national dietary advice bodies (e.g. the UK dietary guidelines^2^) have in recent years recommended the reduction of red and processed meat consumption, based on consistent evidence linking high processed meat, and probably red meat consumption with colorectal cancer risk^1^. While the association between meat intake and cancer risk has been comprehensively studied^3,4^ there is little information on the association between meat consumption, especially poultry meat, and incidence of major non-cancerous health outcomes^5^. This lack of evidence might relate to outcome selection bias (i.e. only reporting the outcomes that are found to be associated with meat^6^), differences in the definition of outcomes, sample size, control of confounders and/or length of follow-up used among different studies. Examining the association between meat consumption and multiple non-cancerous health outcomes in the same cohort may help to clarify these associations^7^.

This study uses an outcome-wide approach to prospectively examine associations of meat consumption with risk of 25 common conditions identified as the 25 leading causes of hospital admission (other than cancer) in a large UK cohort.

## Methods

### Study population

We used data from the UK Biobank study, a cohort of 503 317 men and women from across the UK^8^. Potential participants were recruited through the National Health Service (NHS) Patient Registers and invited to attend one of the 22 assessment centres between 2006 and 2010. Participants joining the study completed a baseline touchscreen questionnaire, provided anthropometric and biological data, and gave informed consent for their health to be followed-up through linkage to electronic medical records. The UK Biobank study was approved by the North West Multi-Centre Research Ethics Committee (reference number 06/MRE08/65).

### Assessment of dietary intake

Dietary intake was assessed using a touchscreen dietary questionnaire administered to all participants at baseline that included 29 questions on diet, assessing the consumption frequency of each listed food. Responses to the five questions on meat (unprocessed beef, unprocessed lamb/mutton, unprocessed pork, unprocessed poultry, and processed meat) were assigned values for frequency per week (never=0, less than once per week = 0.5, once per week=1, 2-4 times per week=3, 5-6 times per week=5.5, and once or more a day=7). We then categorized these meat intake frequencies into three or four categories to create approximately equal-sized groups (see Supplementary Methods S1 for additional detail).

Participants recruited after 2009, as well as participants who provided UK Biobank with an email address and agreed to be re-contacted, additionally filled out the Oxford WebQ^9^, an online 24-hour recall questionnaire. Participants were asked to select how many portions of each food item they consumed over the previous 24-hours, enabling calculation of mean grams per/day by multiplying frequencies of consumption by standard portion sizes. Similar foods were then grouped together into meat types to match the touchscreen dietary questionnaire. We then assigned the mean WebQ intakes in participants who had completed at least three WebQs to each touchscreen category (see Supplementary Methods S1 for additional detail).

### Assessment of health outcomes

The main outcomes of interest in this study were incident cases of 25 common conditions. The conditions selected were those identified as the 25 leading, well-defined causes of non-cancerous hospital admission in this cohort based on the primary International Classification of Diseases (ICD) 10 diagnosis codes recorded during admission. Some of commonest causes of hospital admission in this cohort (e.g. nausea or heartburn) were not considered to be separate conditions, because they were not well-defined and/or were likely to be associated with a diverse range of underlying conditions. Moreover, although diabetes was not among the 25 most common primary diagnoses associated with admission, it is a common secondary reason for admission and therefore *any* diagnosis of diabetes was included among the 25 common conditions examined. (See supplementary Table S1 for selected conditions and relevant diagnosis and procedure codes.)

Participant information on cause-specific in-patient hospital admissions and deaths (primary cause for all outcomes except diabetes which also included underlying cause) was obtained through linkage to the NHS Central Registers. For participants in England, Hospital Episode Statistics (HES) and information on date and cause of death were available until the 31st of March 2017; for participants in Scotland, the Scottish Morbidity Records and information on date and cause of death were available until the 31st of October 2016; and for participants in Wales, the Patient Episode Database and information on date and cause of death were available until the 29th of February 2016. We also obtained information on cancer registrations (including date and cancer site) from the NHS Central Registers. (See Supplementary Methods S2 and Supplementary Table S1 for information on exclusion, diagnosis and procedure codes).

### Exclusions

Of the 503 317 recruited participants, 28 319 were excluded due to study withdrawals, prevalent cancer (except non-melanoma skin cancer, ICD-10 C44) prior to recruitment, or because their genetic sex differed from their reported gender, resulting in a maximal study sample of 474 998 (94%). Participants with a relevant diagnosis or procedure prior to recruitment, ascertained through the touchscreen questionnaire, nurse-guided interviews, and hospital admission data were excluded for each respective analysis (see Supplementary Table S1 for details about the exclusions for each outcome). Participants who did not report their meat intake in the touchscreen questionnaire or reported ‘prefer not to say’ or ‘don’t know’ were classified as missing and excluded for the respective exposure analyses (See Supplementary Figure S1 for participant flowchart and supplementary Tables S6-10 for total numbers for each exposure and outcome).

### Statistical analysis

We used Cox proportional hazards regression models to assess associations between meat consumption and risk for incident cases separately for each disease or condition, calculating trends using the mean meat intakes calculated using the WebQ questionnaires for each category from the touchscreen questionnaire and the trend test variables. Participants’ survival time in person-years was calculated from their age at recruitment until their age at hospital admission, death, loss to follow up, or administrative censoring. All analyses were stratified by sex, age at recruitment, and geographical region (Model 0). In Model 1, we estimated hazard ratios (HRs) and 95% confidence intervals (CIs) adjusted for ethnicity, Townsend deprivation index^10^, education, employment, smoking, alcohol consumption, and physical activity, and in women we additionally adjusted for menopausal status, hormone-replacement therapy, oral contraceptive pill use and parity. In Model 2, we further adjusted for total fruit and vegetable intake and cereal fibre intake score (calculated by multiplying the frequency of consumption of bread and breakfast cereal by the fibre content of these foods^11^). For Model 3, we added adjustment for body mass index (BMI). Missing data for all covariates was minimal (<10%) and thus a ‘missing’ category was created for each covariate. See Figures 1-4 footnotes and Supplementary Methods S3, for full adjustment description with definitions of categories.

**Figure 1.**
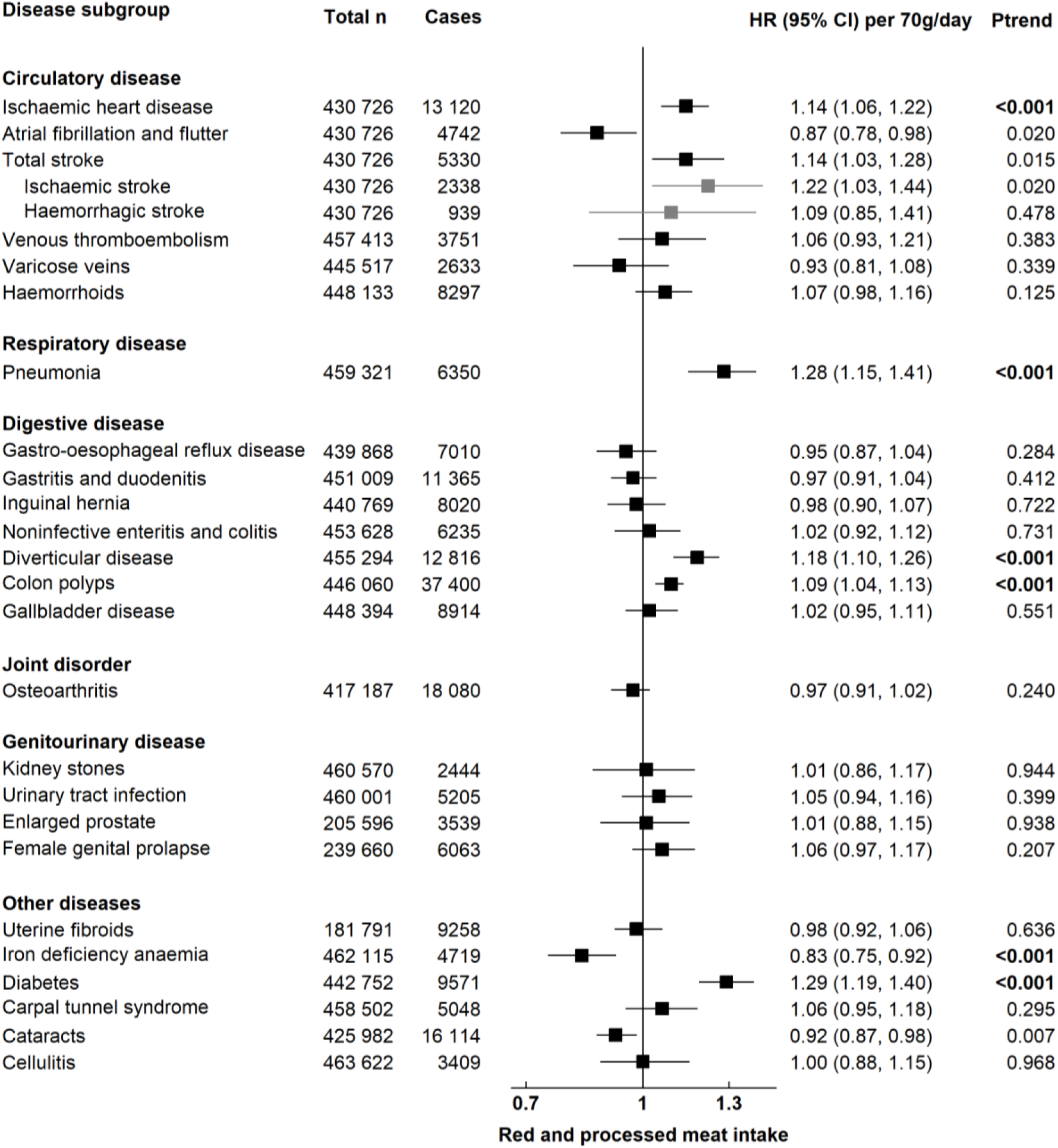
Risk of 25 common conditions per 70 grams higher daily intake of <u>red and processed meat</u>. Stratified for sex, age group and region and adjusted for age (underlying time variable), ethnicity (4 groups where possible: White, Asian or Asian British, Black or Black British, Mixed race or other, unknown), deprivation (Townsend index quintiles, unknown), qualification (College or university degree/vocational qualification, National examination at ages 17-18, National examination at age 16, other/unknown), employment (in paid employment, receiving pension, not in paid employment, unknown), smoking (never, former, current <15 cigarettes/day, current ≥ 15 cigarettes/ day, current unknown amount of cigarettes/day, unknown), physical activity (<10 excess METs per/week, 10-<50 excess METs per/week, ≥ 50 excess METs per/week, unknown), alcohol intake (none, < 1 g/day, 1-<10 g/day, 10-<20 g/day, ≥20 g/day, unknown), total fruit and vegetable intake (< 3 servings/day, 3-< 4 servings/day, 4-< 6 servings/day, ≥ 6 servings/day, unknown), cereal fibre score (sex-specific quintiles, unknown), BMI (sex-specific quintiles, unknown), in women: menopausal status (pre-, postmenopausal, unknown), HRT (never, past, current, unknown), OCP use (never, past, current, unknown), and parity (nulliparous, 1-2, ≥ 3, unknown). BMI: Body mass index, HRT: hormone replacement therapy, OCP: oral contraceptive pill. P trend in bold: P value robust to Bonferroni correction (*P*<0.002).

**Figure 2.**
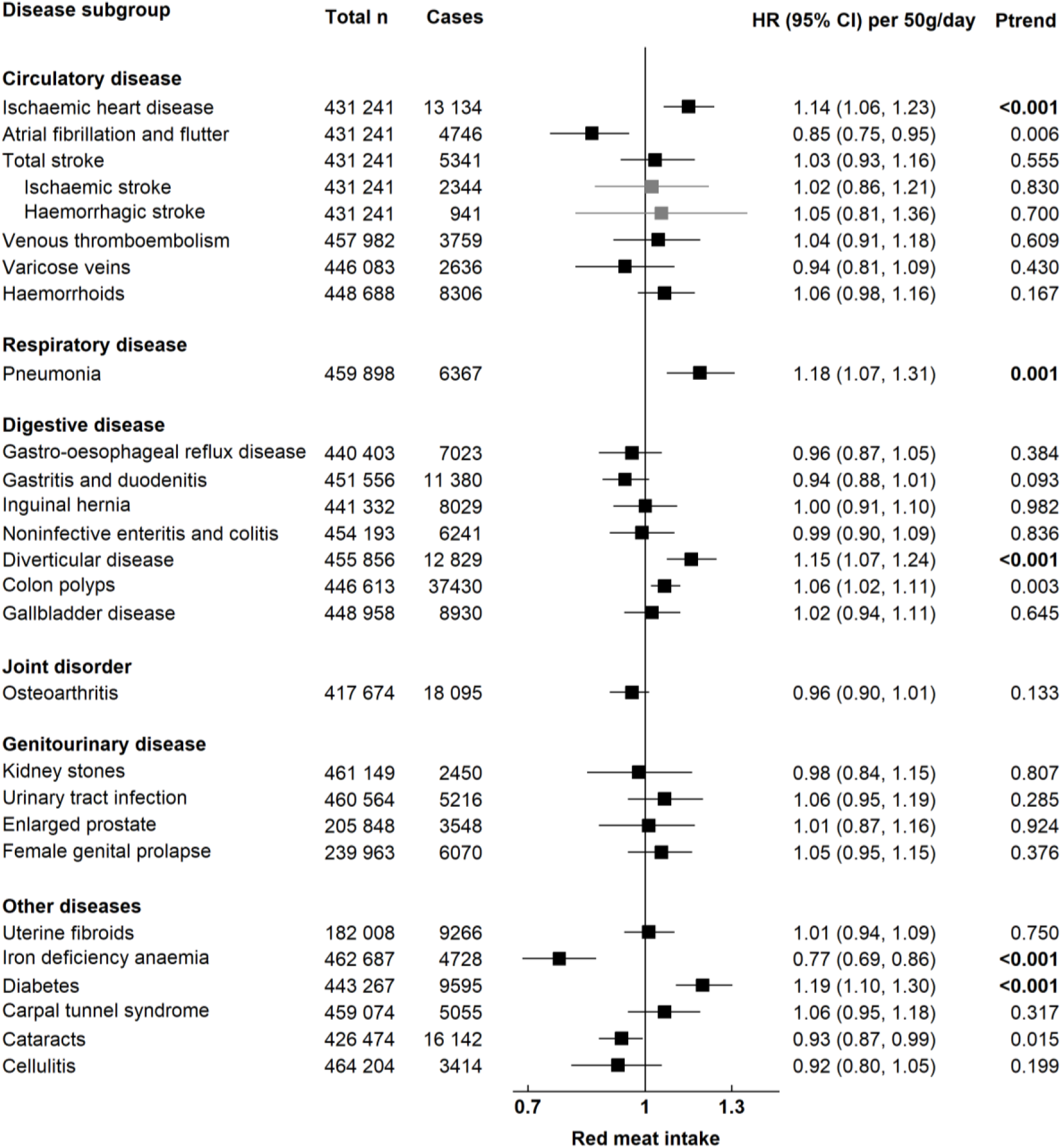
Risk of 25 common conditions per 50 grams higher daily intake of <u>red meat</u>. Stratified for sex, age group and region and adjusted for age (underlying time variable), ethnicity (4 groups where possible: White, Asian or Asian British, Black or Black British, Mixed race or other, unknown), deprivation (Townsend index quintiles, unknown), qualification (College or university degree/vocational qualification, National examination at ages 17-18, National examination at age 16, other/unknown), employment (in paid employment, receiving pension, not in paid employment, unknown), smoking (never, former, current <15 cigarettes/day, current ≥ 15 cigarettes/ day, current unknown amount of cigarettes/day, unknown), physical activity (<10 excess METs per/week, 10-<50 excess METs per/week, ≥ 50 excess METs per/week, unknown), alcohol intake (none, < 1 g/day, 1-<10 g/day, 10-<20 g/day, ≥20 g/day, unknown), total fruit and vegetable intake (< 3 servings/day, 3-< 4 servings/day, 4-< 6 servings/day, ≥ 6 servings/day, unknown), cereal fibre score (sex-specific quintiles, unknown), BMI (sex-specific quintiles, unknown), in women: menopausal status (pre-, postmenopausal, unknown), HRT (never, past, current, unknown), OCP use (never, past, current, unknown), and parity (nulliparous, 1-2, ≥ 3, unknown). BMI: Body mass index, HRT: hormone replacement therapy, OCP: oral contraceptive pill. P trend in bold: P value robust to Bonferroni correction (*P*<0.002).

**Figure 3.**
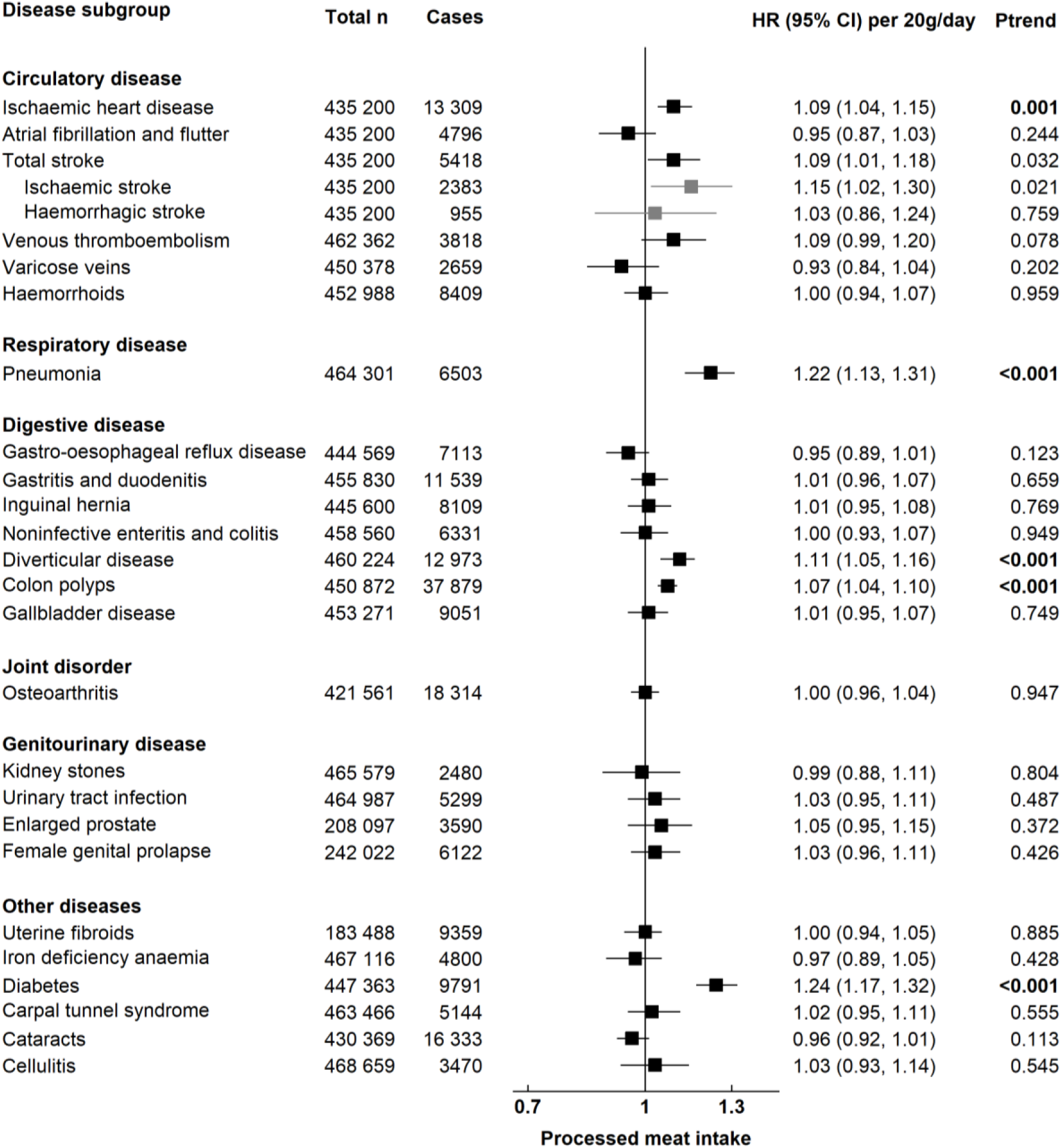
Risk of 25 common conditions per 20 grams higher daily intake of <u>processed meat</u>. Stratified for sex, age group and region and adjusted for age (underlying time variable), ethnicity (4 groups where possible: White, Asian or Asian British, Black or Black British, Mixed race or other, unknown), deprivation (Townsend index quintiles, unknown), qualification (College or university degree/vocational qualification, National examination at ages 17-18, National examination at age 16, other/unknown), employment (in paid employment, receiving pension, not in paid employment, unknown), smoking (never, former, current <15 cigarettes/day, current ≥ 15 cigarettes/ day, current unknown amount of cigarettes/day, unknown), physical activity (<10 excess METs per/week, 10-<50 excess METs per/week, ≥ 50 excess METs per/week, unknown), alcohol intake (none, < 1 g/day, 1-<10 g/day, 10-<20 g/day, ≥20 g/day, unknown), total fruit and vegetable intake (< 3 servings/day, 3-< 4 servings/day, 4-< 6 servings/day, ≥ 6 servings/day, unknown), cereal fibre score (sex-specific quintiles, unknown), BMI (sex-specific quintiles, unknown), in women: menopausal status (pre-, postmenopausal, unknown), HRT (never, past, current, unknown), OCP use (never, past, current, unknown), and parity (nulliparous, 1-2, ≥ 3, unknown). BMI: Body mass index, HRT: hormone replacement therapy, OCP: oral contraceptive pill. P trend in bold: P value robust to Bonferroni correction (*P*<0.002).

**Figure 4.**
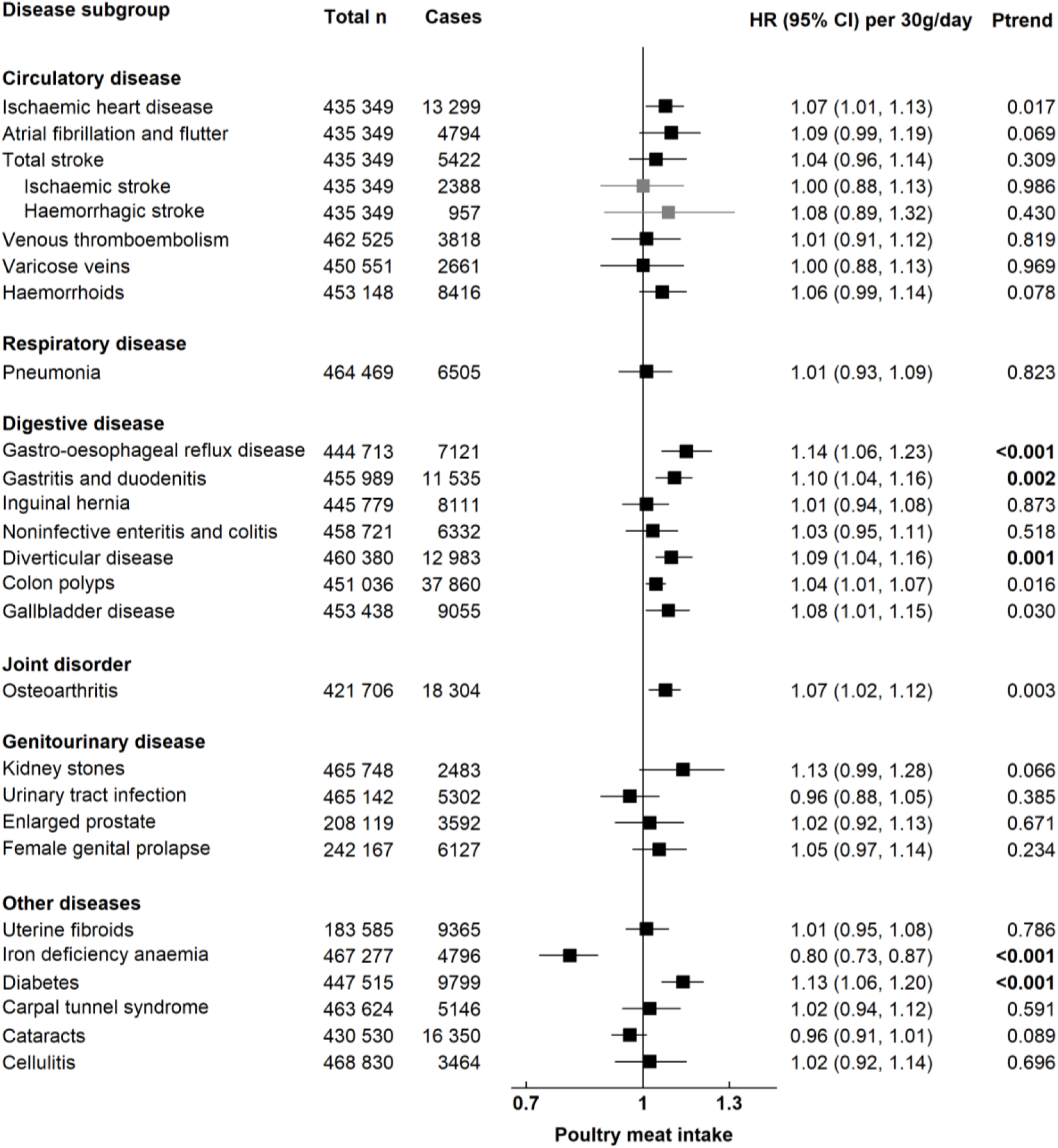
Risk of 25 common conditions per 30 grams higher daily intake of <u>poultry meat</u>. Stratified for sex, age group and region and adjusted for age (underlying time variable), ethnicity (4 groups where possible: White, Asian or Asian British, Black or Black British, Mixed race or other, unknown), deprivation (Townsend index quintiles, unknown), qualification (College or university degree/vocational qualification, National examination at ages 17-18, National examination at age 16, other/unknown), employment (in paid employment, receiving pension, not in paid employment, unknown), smoking (never, former, current <15 cigarettes/day, current > 15 cigarettes/ day, current unknown amount of cigarettes/day, unknown), physical activity (<10 excess METs per/week, 10-<50 excess METs per/week, ≥ 50 excess METs per/week, unknown), alcohol intake (none, < 1 g/day, 1-<10 g/day, 10-<20 g/day, ≥20 g/day, unknown), total fruit and vegetable intake (< 3 servings/day, 3-< 4 servings/day, 4-< 6 servings/day, ≥ 6 servings/day, unknown), cereal fibre score (sex-specific quintiles, unknown), BMI (sex-specific quintiles, unknown), in women: menopausal status (pre-, postmenopausal, unknown), HRT (never, past, current, unknown), OCP use (never, past, current, unknown), and parity (nulliparous, 1-2, ≥ 3, unknown). BMI: Body mass index, HRT: hormone replacement therapy, OCP: oral contraceptive pill. P trend in bold: P value robust to Bonferroni correction (P<0.002).

#### Sensitivity analyses

To examine whether the associations between meat intake and risk of incidence for specific diagnoses could be affected by reverse causality or residual confounding by smoking, we repeated the analyses after excluding the first 4 years of follow-up and only in never smokers.

All analyses were conducted using STATA version 15.1 (Stata Corp LP, College Station, TX). All *P* values were two-sided and Bonferroni correction was used to allow for multiple testing (for 25 outcomes, *P*<0.002).

## Results

### Baseline characteristics

Table 1 shows baseline characteristics of participants by categories of red and processed meat intake. Around one-third of participants consumed red and/or processed meat once or more daily. On average, participants who consumed red and processed meat regularly (three or more times per week) were more likely to be men, older, of White European ethnicity, retired, have higher BMI, smoke and consume alcohol, consume less fruit and fibre and more poultry meat; they were also less likely to have attained a tertiary education, and among women to have three or more children, use oral contraceptives or hormone replacement therapy, or be postmenopausal compared with participants who consumed meat less than three times per week (*P* <0.001 for heterogeneity between meat intakes for all baseline characteristics). Participants who consumed higher amounts of red meat were more likely to consume higher amounts of processed and poultry meat (see Supplementary Table S3). The characteristics in relation to poultry meat consumption were somewhat different (see Supplementary Table S5).

**Table 1.** Baseline characteristics of participants by red and processed meat intake in UK Biobank (n=467,754^a^)

| Characteristic | 0-1 times/week<br>N=44 021 | 2 times/week<br>N=160 073 | 3-4 times/week<br>N=140 678 | ≥ 5times/week<br>N=122 982 |
| --- | --- | --- | --- | --- |
| **Sociodemographic** |  |  |  |  |
| Sex, n (%) |  |  |  |  |
| Women | 31 319 (71.1) | 101 750 (63.6) | 71 681 (51.0) | 47 657 (38.8) |
| Men | 12 702 (28.9) | 58 323 (36.4) | 68 997 (49.0) | 75 325 (61.2) |
| Age (years), mean (SD) | 54.9 (8.2) | 56.5 (8.0) | 56.4 (8.1) | 56.5 (8.2) |
| Ethnicity, n (%) |  |  |  |  |
| White | 38 453 (87.4) | 151 731 (94.8) | 134 513 (95.6) | 116 818 (95.0) |
| Asian or Asian British | 3 468 (7.9) | 2 914 (1.8) | 2 245 (1.6) | 1 897 (1.5) |
| Black or Black British | 886 (2.0) | 2 607 (1.6) | 1 685 (1.2) | 2 047 (1.7) |
| Mixed Race/Other | 997 (2.3) | 2 323 (1.5) | 1 807 (1.3) | 1 774 (1.4) |
| Unknown | 217 (0.5) | 498 (0.3) | 428 (0.3) | 446 (0.4) |
| Townsend deprivation, n (%) |  |  |  |  |
| Most affluent (mean -4.7) | 7 037 (16.0) | 32 848 (20.5) | 29 803 (21.2) | 24 666 (20.1) |
| 2 (mean -3.3) | 7 525 (17.1) | 32 746 (20.5) | 29 079 (20.7) | 24 304 (19.8) |
| 3 (mean -2.1) | 8 341 (18.9) | 32 738 (20.5) | 28 325 (20.1) | 24 343 (19.8) |
| 4 (mean -0.1) | 10 146 (23.0) | 31 755 (19.8) | 27 447 (19.5) | 24 069 (19.6) |
| Most deprived (mean 3.8) | 10 910 (24.8) | 29 771 (18.6) | 25 871 (18.4) | 25 448 (20.7) |
| Unknown | 62 (0.1) | 215 (0.1) | 153 (0.1) | 152 (0.1) |
| Qualification, n (%) |  |  |  |  |
| College/university degree/NVQ | 28 492 (64.7) | 96 077 (60.0) | 82 367 (58.6) | 72 438 (58.9) |
| National examination at ages 17-18 | 2 491 (5.7) | 8 708 (5.4) | 7 795 (5.5) | 6 626 (5.4) |
| National examination at age 16 | 6 321 (14.4) | 27 682 (17.3) | 24 134 (17.2) | 19 676 (16.0) |
| Other/unknown | 6 717 (15.3) | 27 606 (17.2) | 26 382 (18.8) | 24 242 (19.7) |
| Employment, n (%) |  |  |  |  |
| In paid employment | 27 652 (62.8) | 93 968 (58.7) | 81 645 (58.0) | 70 102 (57.0) |
| Pension | 10 229 (23.2) | 48 220 (30.1) | 42 698 (30.4) | 37 181 (30.2) |
| Not in paid employment | 5 556 (12.6) | 16 487 (10.3) | 15 236 (10.8) | 14 563 (11.8) |
| Unknown | 584 (1.3) | 1 398 (0.9) | 1 099 (0.8) | 1 136 (0.9) |
| **Physical measurements** |  |  |  |  |
| BMI (kg/m<sup>2</sup>), mean (SD) | 25.9 (4.7) | 27.1 (4.7) | 27.6 (4.8) | 28.1 (4.9) |
| **Lifestyle** |  |  |  |  |
| Smoking, n (%) |  |  |  |  |
| None | 26 347 (59.9) | 90 449 (56.5) | 76 686 (54.5) | 62 688 (51.0) |
| Former | 13 964 (31.7) | 54 799 (34.2) | 48 534 (34.5) | 43 270 (35.2) |
| Current <15 cigarettes/day | 1 297 (2.9) | 4 581 (2.9) | 4 247 (3.0) | 4 074 (3.3) |
| Current ≥15 cigarettes/day | 1 030 (2.3) | 4 825 (3.0) | 6 015 (4.3) | 7 623 (6.2) |
| Current, amount unknown | 1 213 (2.8) | 4 876 (3.0) | 4 727 (3.4) | 4 914 (4.0) |
| Unknown | 170 (0.4) | 543 (0.3) | 469 (0.3) | 413 (0.3) |
| Physical activity level, n (%) |  |  |  |  |
| Low <10 excess METs | 7 770 (17.7) | 24 716 (15.4) | 21 860 (15.5) | 20 458 (16.6) |
| Moderate 10-<50 excess METs | 22 380 (50.8) | 80 041 (50.0) | 68 197 (48.5) | 58 007 (47.2) |
| High ≥ 50 excess METs | 12 406 (28.2) | 49 710 (31.1) | 45 470 (32.3) | 39 708 (32.3) |
| Unknown | 1 465 (3.3) | 5 606 (3.5) | 5 151 (3.7) | 4 809 (3.9) |
| Alcohol intake, n (%) |  |  |  |  |
| non-drinkers | 6 814 (15.5) | 19 867 (12.4) | 14 469 (10.3) | 10 862 (8.8) |
| <1g/d | 14 743 (33.5) | 57 212 (35.7) | 43 228 (30.7) | 31 588 (25.7) |
| 1-<10g/d | 7 985 (18.1) | 36 199 (22.6) | 31 682 (22.5) | 25 627 (20.8) |
| 10-<20g/d | 6 740 (15.3) | 34 046 (21.3) | 41 069 (29.2) | 46 545 (37.8) |
| 20+g/d | 7 503 (17.0) | 11 939 (7.5) | 9 611 (6.8) | 7 791 (6.3) |
| Unknown | 236 (0.5) | 810 (0.5) | 619 (0.4) | 569 (0.5) |
| **Diet** |  |  |  |  |
| Fruit & vegetable intake (s/day), mean (SD) | 5.59 (3.19) | 4.89 (2.54) | 4.50 (2.45) | 4.33 (2.50) |
| Fruit & vegetable intake categories, n (%) |  |  |  |  |
| <3 servings/day | 5 486 (12.5) | 25 992 (16.2) | 29 853 (21.2) | 29 895 (24.3) |
| 3-<4 servings/day | 6 014 (13.7) | 27 540 (17.2) | 27 245 (19.4) | 25 094 (20.4) |
| 4-<6 servings/day | 14 692 (33.4) | 58 168 (36.3) | 48 946 (34.8) | 40 319 (32.8) |
| ≥6 servings/day | 16 938 (38.5) | 45 648 (28.5) | 32 163 (22.9) | 25 227 (20.5) |
| Unknown | 891 (2.0) | 2 725 (1.7) | 2 471 (1.8) | 2 447 (2.0) |
| Cereal fibre intake (g), mean (SD) | 4.66 (3.11) | 4.52 (2.89) | 4.52 (2.91) | 4.44 (2.96) |
| Poultry meat, n (%) |  |  |  |  |
| 0-1 times/week | 26 361 (59.9) | 23 488 (14.7) | 13 478 (9.6) | 10 760 (8.7) |
| 2 times/week | 7 141 (16.2) | 61 115 (38.2) | 56 697 (40.3) | 42 293 (34.4) |
| ≥3 times/week | 10 461 (23.8) | 75 360 (47.1) | 70 407 (50.0) | 69 836 (56.8) |
| Unknown | 58 (0.1) | 110 (0.1) | 96 (0.1) | 93 (0.1) |
| Menopausal status, n (%) |  |  |  |  |
| Premenopausal | 8 960 (20.4) | 22 947 (14.3) | 17 039 (12.1) | 11 207 (9.1) |
| Postmenopausal | 20 589 (46.8) | 73 269 (45.8) | 50 661 (36.0) | 33 725 (27.4) |
| Unknown | 1 770 (4.0) | 5 534 (3.5) | 3 981 (2.8) | 2 725 (2.2) |
| Parity, n (%) |  |  |  |  |
| 0 births | 8 314 (18.9) | 19 782 (12.4) | 11 739 (8.3) | 7 298 (5.9) |
| 1 - 2 births | 16 232 (36.9) | 58 206 (36.4) | 42 076 (29.9) | 27 654 (22.5) |
| ≥3 births | 6 717 (15.3) | 23 671 (14.8) | 17 812 (12.7) | 12 655 (10.3) |
| Unknown | 56 (0.1) | 91 (0.1) | 54 (0.0) | 50 (0.0) |
| HRT use, n (%) |  |  |  |  |
| Never | 21 437 (48.7) | 61 913 (38.7) | 44 008 (31.3) | 28 975 (23.6) |
| Past | 7 951 (18.1) | 33 163 (20.7) | 23 018 (16.4) | 15 560 (12.7) |
| Current | 1 773 (4.0) | 6 379 (4.0) | 4 429 (3.1) | 2 897 (2.4) |
| Unknown | 158 (0.4) | 295 (0.2) | 226 (0.2) | 225 (0.2) |
| OCP use, n (%) |  |  |  |  |
| Never | 6 596 (15.0) | 18 181 (11.4) | 12 850 (9.1) | 8 917 (7.3) |
| Past | 23 875 (54.2) | 81 529 (50.9) | 57 267 (40.7) | 37 642 (30.6) |
| Current | 691 (1.6) | 1 813 (1.1) | 1 388 (1.0) | 918 (0.7) |
| Unknown | 157 (0.4) | 227 (0.1) | 176 (0.1) | 180 (0.1) |

### Risk analyses

Figures 1-4 present the numbers of incident cases for 25 common conditions and their HRs and 95% CIs per unit higher intake of meat for the multiple-adjusted model over an average follow-up of 8.0 years (standard deviation 1.0). Risks by categories of meat intake at baseline for Models 0-3 can be found in Supplementary Tables S6-10. Overall, many of the positive, associations were attenuated, and in some cases became null with the additional adjustment for BMI (Model 3). Here we describe the results for Model 3 that were robust to correction for multiple testing.

#### Red and processed meat

Red and processed meat intake was associated with a higher risk of ischaemic heart disease (IHD) (HR per 70 g/day higher intake =1.14, 95% CI 1.06-1.22), pneumonia (1.28, 1.15-1.41), diverticular disease (1.18, 1.10-1.26), colon polyps (1.09, 1.04-1.13) and diabetes (1.29, 1.19-1.40), and a lower risk of iron deficiency anaemia (IDA) (0.83, 0.75-0.92) (Figure 1).

#### Red meat

Red meat intake was associated with a higher risk of IHD (HR per 50 g/day higher intake =1.14, 95% CI 1.06, 1.23), pneumonia (1.18, 1.07-1.31), diverticular disease (1.15, 1.07-1.24) and diabetes (1.19, 1.10-1.30), and a lower risk of IDA (0.77, 0.69-0.86) (Figure 2).

#### Processed meat

Processed meat intake was associated with a higher risk of IHD (HR per 20 g/day higher intake =1.09, 95% CI 1.04-1.15), pneumonia (1.22, 95% CI 1.13-1.31), diverticular disease (1.11, 1.05-1.16,) colon polyps (1.07, 95% CI 1.04-1.10) and diabetes (1.24, 1.17-1.32) (Figure 3).

#### Poultry meat

Poultry meat intake was associated with a higher risk of diverticular disease (HR per 30 g/day higher intake =1.09, 95% CI 1.04-1.16), gastro-oesophageal reflux disease (GERD) (1.14, 1.06-1.23), gastritis and duodenitis (1.10, 1.04-1.16), and diabetes (1.13, 1.06-1.20), and a lower risk of IDA (0.80, 0.73-0.87) (Figure 4).

#### Sensitivity analysis

Associations were similar when excluding the first 4 years of follow-up and in never smokers (Supplementary Figures S2-6). However, we did note a positive association between red and processed meat intake (combined) and haemorrhagic stroke (HR per 70 g/day higher intake =1.49, 95% CI 1.07-2.08) in participants diagnosed after four or more years of follow-up; and that the associations between red meat intake and diabetes risk, and processed meat intake and IHD risk, were null in never smokers.

## Discussion

In this large, prospective cohort of nearly 0.5 million UK adults, we observed that after allowing for multiple testing higher consumption of red and processed meat combined was associated with higher risks of IHD, pneumonia, diverticular disease, colon polyps, and diabetes, and higher consumption of poultry meat was associated with higher risks of GERD, gastritis and duodenitis, diverticular disease, and diabetes. Differences in BMI across the categories of meat consumption appear to account for some of the increased risks. We also observed inverse associations between higher intakes of red and poultry meat and IDA, which were minimally affected by adjustment for BMI.

### Circulatory diseases

Similar to our findings, a recent meta-analysis of prospective studies^12^ and a recent prospective study from the Pan-European EPIC cohort which included over 7000 IHD cases^13^ reported positive associations between red meat and processed meat consumption and risk of IHD. For stroke, previous meta-analyses of prospective studies^14,15^ and a recent prospective study from the EPIC cohort^16^ both reported null associations for red and processed meat intake and haemorrhagic stroke; this is consistent with our main findings but not with our findings in participants diagnosed after four or more years of follow-up; though this might be a chance finding due to shorter follow-up. Processed meats contain high amounts of sodium^17^, a risk factor for high blood pressure^18^, which is a causal risk factor for IHD and stroke^19^. Furthermore, processed meat is a major dietary source of saturated fatty acids (SFAs) which can increase low-density lipoprotein (LDL) cholesterol, an established causal risk factor for IHD^20^.

### Respiratory disease

Higher consumption of red and processed meat was associated with a higher risk of pneumonia; to the best of our knowledge these associations have not been shown previously. It is possible that hospital admission for pneumonia is a marker for co-morbidity and overall frailty^21^, therefore residual confounding might operate (see further discussion on residual confounding below). It is also possible that the observed association might reflect a causal link, for example related to the high availability of iron in red and processed meat (see further discussion below in relation to anaemia). Excess iron has been found to be associated with a higher risk of infection^22^; and increased availability of iron for invading bacterial species and other pathogens^23^.

### Digestive diseases

Few prospective studies have examined risk for diverticular disease^24,25^, but consistent with our findings the Health Professionals Follow-up Study (HPFS) observed increased risks of incident diverticulitis with higher consumption of red and processed meat^24^. The HPFS did not observe an association for poultry meat, but had much lower power than the current study. Meat consumption might affect the risk of diverticular disease via the microbiome, and there is some evidence that meat intake might alter microbial community structure and change the metabolism of bacteria^26^.

A recent meta-analysis of prospective studies reported that red and processed meat consumption was positively associated with the risk of colorectal adenomas^27^, which is consistent with our findings for colon polyps. Red meat is a source of heme iron and processed meat usually contains nitrite and nitrates; these can promote the formation of N-nitroso compounds^28^, which are mutagenic and have been associated with a higher risk of colorectal adenomas^29^.

To our knowledge, this is the first prospective study of meat consumption and risk of GERD and gastritis and duodenitis. We found a positive association between poultry meat intake and GERD risk, whereas the available cross-sectional evidence suggests a null association for meat (total)^30-33^. We also found a positive association between poultry meat consumption and risk of gastritis and duodenitis. *Helicobacter pylori*, a bacteria that increases the risk of gastritis^34^, has been previously detected in raw poultry meat^35^. Therefore, it is possible that the observed association might relate to inappropriate handling or cooking of poultry meat, but additional research is needed.

### Other diseases

We found an inverse association between the consumption of red and processed meat combined, red meat, and poultry meat and risk of IDA. Some previous evidence from prospective studies^36^ supports these findings, and has also shown a positive association between red meat^37^ and total meat^38-40^ consumption and indicators of body iron stores. Moreover, previous cross-sectional work from the UK Biobank has shown that people who did not consume meat were more likely to be anaemic^41^. This association is likely related to the high availability of heme iron in meat, which is more easily absorbed than non-heme iron (found in plant sources)^42^

Similar to our findings, meta-analyses of prospective cohort studies have consistently reported a positive association between red and processed meat consumption and risk of diabetes^43-45^. We also found a positive association between poultry meat consumption and risk of diabetes, which has been reported in some^46^ but not all prospective studies ^47,48^. The positive association observed for meat consumption and diabetes might relate to heme iron intake and greater iron storage in the body. In high amounts, iron, a pro-oxidant, can promote the formation of hydroxyl radicals that may attack the pancreatic beta cells, thereby impairing insulin synthesis and excretion^49,50^.

### Possible role of BMI

In the present study, most of the positive associations between meat consumption and health risks were substantially attenuated after adjusting for BMI, suggesting that BMI was a strong confounder or possible mediator for many of the meat and disease associations; BMI was highest in participants who consumed meat most frequently, and BMI is an important risk factor for many of the diseases examined (e.g diabetes^45^). Moreover, the associations which remain after adjustment for BMI might still be partly due to residual confounding, because BMI is not a perfect measure of adiposity.

### Strengths and limitations

This is as far as we are aware the first outcome-wide study of meat intake and risk of 25 common conditions (other than cancer). Additional strengths of this study include the large size of the cohort, its prospective design, and wide array of included confounders. This allowed us to investigate a large number of common conditions while simultaneously controlling for confounding, and thus avoid outcome selection bias. Additionally, we used national record linkage to ascertain information on disease incidence. Nevertheless, some potential methodological issues should be considered when interpreting our findings. Some measurement error would have occurred while measuring meat consumption at baseline; however, we reduced the likely effect of random error and short-term variation in diet by using the repeated 24-hour recall WebQ data and applying corrected intakes to each category of the baseline intakes. Additionally, multiple testing might have led to some spurious findings; however, we addressed this by using Bonferroni correction. Another consideration is the use of hospital records for incident case ascertainment. Some conditions might only require hospital use at later stages (e.g. diabetes), and therefore some admissions might reflect prevalent and/or more severe cases. Finally, given the observational nature of this study, it is possible that there is still unmeasured confounding, residual confounding and reverse causality. For instance, in analyses restricted to never smokers, some of the adjusted risk estimates were lower than in the main analysis (e.g. for red meat intake and diabetes and for processed meat intake and IHD), suggesting that even after adjustment for smoking there may be residual confounding. However, most of our results were largely similar after excluding participants who smoked or formerly smoked, and the first four years of follow-up.

## Conclusions

Our findings from this large, prospective study of British adults show that red and poultry meat consumption is associated with a lower risk of IDA. Meat consumption is also associated with higher risks of several common conditions, at least partly accounted for by BMI; and additional research is needed to evaluate whether these differences in risk reflect causal relationships.

## Data Availability

No additional data are available.

## Acknowledgements

This research has been conducted using the UK Biobank Resource under application number 24494. We thank all participants, researchers and support staff who make the study possible.

